# SARS-CoV-2 variant exposures elicit antibody responses with differential cross-neutralization of established and emerging strains including Delta and Omicron

**DOI:** 10.1101/2021.09.08.21263095

**Authors:** Matthew T Laurie, Jamin Liu, Sara Sunshine, James Peng, Douglas Black, Anthea M Mitchell, Sabrina A Mann, Genay Pilarowski, Kelsey C Zorn, Luis Rubio, Sara Bravo, Carina Marquez, Joseph J Sabatino, Kristen Mittl, Maya Petersen, Diane Havlir, Joseph DeRisi

## Abstract

The wide spectrum of SARS-CoV-2 variants with phenotypes impacting transmission and antibody sensitivity necessitates investigation of the immune response to different spike protein versions. Here, we compare the neutralization of variants of concern, including B.1.617.2 (Delta) and B.1.1.529 (Omicron) in sera from individuals exposed to variant infection, vaccination, or both. We demonstrate that neutralizing antibody responses are strongest against variants sharing certain spike mutations with the immunizing exposure. We also observe that exposure to multiple spike variants increases the breadth of variant cross-neutralization. These findings contribute to understanding relationships between exposures and antibody responses and may inform booster vaccination strategies.

**SUMMARY:** This study characterizes neutralization of eight different SARS-CoV-2 variants, including Delta and Omicron, with respect to nine different prior exposures, including vaccination, booster, and infections with Delta, Epsilon, and others. Different exposures were found to confer substantially differing neutralization specificity.

## Background

Genomic surveillance of SARS-CoV-2 continues to identify a diverse spectrum of emerging variants possessing mutations in the spike gene, the main viral determinant of cellular entry and primary target of neutralizing antibodies [1]. Many spike mutations likely result from selective pressure which improves viral fitness through increased transmissibility or evasion of host immunity [2,3]. Studies have demonstrated that sera from vaccinated and naturally infected individuals yield diminished neutralizing activity against certain variants, including the globally dominant Delta variant [4]. Because serum neutralization titer is an important correlate of real-world protective immunity, these findings suggest that antibody responses elicited by exposure to ancestral spike versions (Wuhan or D614G) will be less effective at preventing future infection by certain variants [5]. However, the diversity and prevalence of variants have fluctuated greatly throughout the pandemic, creating a complex population of individuals that may have inherently different capacity to neutralize certain variants depending on the specific genotype of their previous exposures, including vaccination [6].

In this study, we address the question of variant-elicited immune specificity by determining the breadth of neutralizing activity elicited by exposure to specific SARS-CoV-2 variants, vaccines, or both. To accomplish this, we collected serum from subjects with prior infections by variants B.1 (D614G mutation only), B.1.429 (Epsilon), P.2 (Zeta), B.1.1.519, and B.1.617.2 (Delta), which were identified by viral sequencing. We also collected serum from mRNA vaccine recipients who were infected with the B.1 ancestral spike lineage prior to vaccination, infected with B.1.429 prior to vaccination, or had no prior infection. We measured and compared the neutralization titer of each serum cohort against a panel of pseudoviruses representing each different exposure variant plus the variants of concern B.1.351 (Beta), P.1 (Gamma), B.1.617, B.1.617.2 (Delta), and B.1.1.529 (Omicron), which have one or more spike mutations of interest in common with one of the exposure variants. Our results provide a quantitative comparison of the degree of neutralization specificity produced by different exposures. We also demonstrate the effect of serial exposure to different spike versions in broadening the cross-reactivity of neutralizing antibody responses. Together, these findings describe correlates of protective immunity within the rapidly evolving landscape of SARS-CoV-2 variants and are highly relevant to the design of future vaccination strategies targeting spike antigens.

## Methods

### Serum collection

Samples for laboratory studies were obtained under informed consent from participants in an ongoing community program “Unidos en Salud”, which provides SARS-CoV-2 testing, genomic surveillance, and vaccination services in San Francisco, California [7]. Subjects with and without symptoms of COVID-19 were screened with the BinaxNOW rapid antigen assay (supplied by California Department of Public Health). Positive rapid tests were followed by immediate disclosure and outreach to household members for testing, supportive community services, and academic partnership for research studies. All samples were sequenced using ARTIC Network V3 primers on an Illumina NovaSeq platform and consensus genomes generated from the resulting raw .fastq files using IDseq [8].

Convalescent serum donors were selected based on sequence-confirmed infection with the following variants of interest: B.1 (D614G mutation only; n=10 donors), B.1.429 (Epsilon; n=15), B.1.1.519 (n=6), P.2 (Zeta; n=1), B.1.526 (Iota; n=1), B.1.617.2 (Delta; n=3), D614G infection with subsequent BNT162b2 vaccination (n=8), and B.1.429 infection with subsequent BNT162b2 vaccination (n=17). Serum was also collected from healthy recipients of two (n=11) or three (n=7) doses of BNT162b2 or mRNA-1273 vaccines who were confirmed to have no prior SARS-CoV-2 infection by anti-SARS-CoV-2 nucleocapsid IgG assay [9]. All serum was collected from donors an average of 34 days (standard deviation 16.6 days) after exposure to either SARS-CoV-2 or the most recent dose of mRNA vaccine. For pooled serum experiments, samples from the same exposure group were pooled at equal volumes. Serum samples from the closely related exposures P.2 and B.1.526 were pooled together for the “E484K exposure” pool, and samples from BNT162b2 and mRNA-1273 exposures were pooled together for the “vaccine exposure” pool because of the very similar neutralization specificity observed in individual tests of these sera. Serum samples were heat inactivated at 56°C for 30 minutes prior to experimentation. Relevant serum sample metadata and exposure grouping is shown in Table S1A.

### Pseudovirus production

SARS-CoV-2 pseudoviruses bearing spike proteins of variants of interest were generated using a recombinant vesicular stomatitis virus expressing GFP in place of the VSV glycoprotein (rVSVΔG-GFP) described previously [10]. The following mutations representative of specific spike variants were cloned in a CMV-driven expression vector and used to produce SARS-CoV-2 spike pseudoviruses: B.1 (D614G), B.1.429/Epsilon (S13I, W152C, L452R, D614G), P.2/Zeta (E484K, D614G), B.1.351/Beta (D80A, D215G, Δ242-244, K417N, E484K, N501Y, D614G, A701V), P.1/Gamma (L18F, T20N, P26S, D138Y, R190S, K417T, E484K, N501Y, D614G, H655Y, T1027I, V1176F), B.1.1.519 (T478K, D614G, P681H, T732A), B.1.617 (L452R, E484Q, D614G, P681R), B.1.617.2/Delta (T19R, T95I, G142D, Δ157-158, L452R, T478K, P681R, D614G, D950N), and B.1.1.529/Omicron (32 spike mutations). All pseudovirus spike mutations are listed in Table S1C. Pseudoviruses were titered on Huh7.5.1 cells overexpressing ACE2 and TMPRSS2 (gift of Andreas Puschnik) using GFP expression to measure the concentration of focus forming units (ffu).

### Pseudovirus neutralization experiments

Huh7.5.1-ACE2-TMPRSS2 cells were seeded in 96-well plates at a density of 7000 cells/well one day prior to pseudovirus inoculation. Serum samples were diluted into complete culture media (DMEM with 10% FBS, 10mM HEPES, 1x Pen-Strep-Glutamine) using the LabCyte Echo 525 liquid handler and 1500 ffu of each pseudovirus was added to the diluted serum to reach final dilutions ranging from 1:40-1:5120, including no-serum and no-pseudovirus controls. Serum/pseudovirus mixtures were incubated at 37°C for 1h before being added directly to cells. Cells inoculated with serum/pseudovirus mixtures were incubated at 37°C and 5% CO_2_ for 24h, resuspended using 10x TrypLE Select (Gibco), and cells were assessed with the BD Celesta flow cytometer. The WHO International Reference Standard 20/150 was used to validate the pseudovirus assay and compare serum neutralization titers (Table S1B) [11]. All neutralization assays were repeated in a total of three independent experiments with each experiment containing two technical replicates for each condition. Cells were verified to be free of mycoplasma contamination with the MycoAlert Mycoplasma detection kit (Lonza).

### Data analysis

Pseudovirus flow cytometry data was analyzed with FlowJo to determine the percentage of GFP-positive cells, indicating pseudovirus transduction. Percent neutralization for each condition was calculated by normalizing GFP-positive cell percentage to no-serum control wells. Neutralization titers (NT_50_ and NT_90_) were calculated from eight-point response curves generated in GraphPad Prism 7 using four-parameter logistic regression. The fold-change in pseudovirus neutralization titer in each serum group was calculated by normalizing each variant NT_50_ and NT_90_ value to D614G pseudovirus NT_50_ and NT_90_ values in the same serum group. To compare neutralization titer across a panel of different pseudoviruses and serum groups, the Log2 fold-change compared to D614G pseudovirus was reported.

## Results

We compared the 50% and 90% neutralization titers (NT_50_ and NT_90_) of D614G and B.1.429 (Epsilon) pseudoviruses in individual serum samples from subjects exposed to D614G infection, B.1.429 infection, mRNA vaccination, D614G infection with subsequent mRNA vaccination, and B.1.429 infection with subsequent mRNA vaccination (Figure 1). Fold-changes in both NT_50_ and NT_90_ are reported since these values often differ in magnitude due to differences in neutralization curve slope between different variants and sera. In D614G-exposed and vaccine-exposed serum, we observed approximately 2 to 3-fold decreases in average neutralization titer against B.1.429 pseudovirus compared to D614G pseudovirus. As expected, B.1.429-exposed serum neutralized B.1.429 pseudovirus more efficiently than D614G pseudovirus. Of note, previous infection with either D614G or B.1.429 followed by vaccination led to substantially higher neutralization titers against both pseudoviruses. In contrast to other exposure groups, serum from vaccine recipients previously infected by B.1.429 neutralized D614G and B.1.429 at similar titers, with only a 1.3-fold difference in NT_90_, indicating that exposure to multiple spike variants elicits a potent response with specificity toward the breadth of prior exposures.

**Figure 1.**
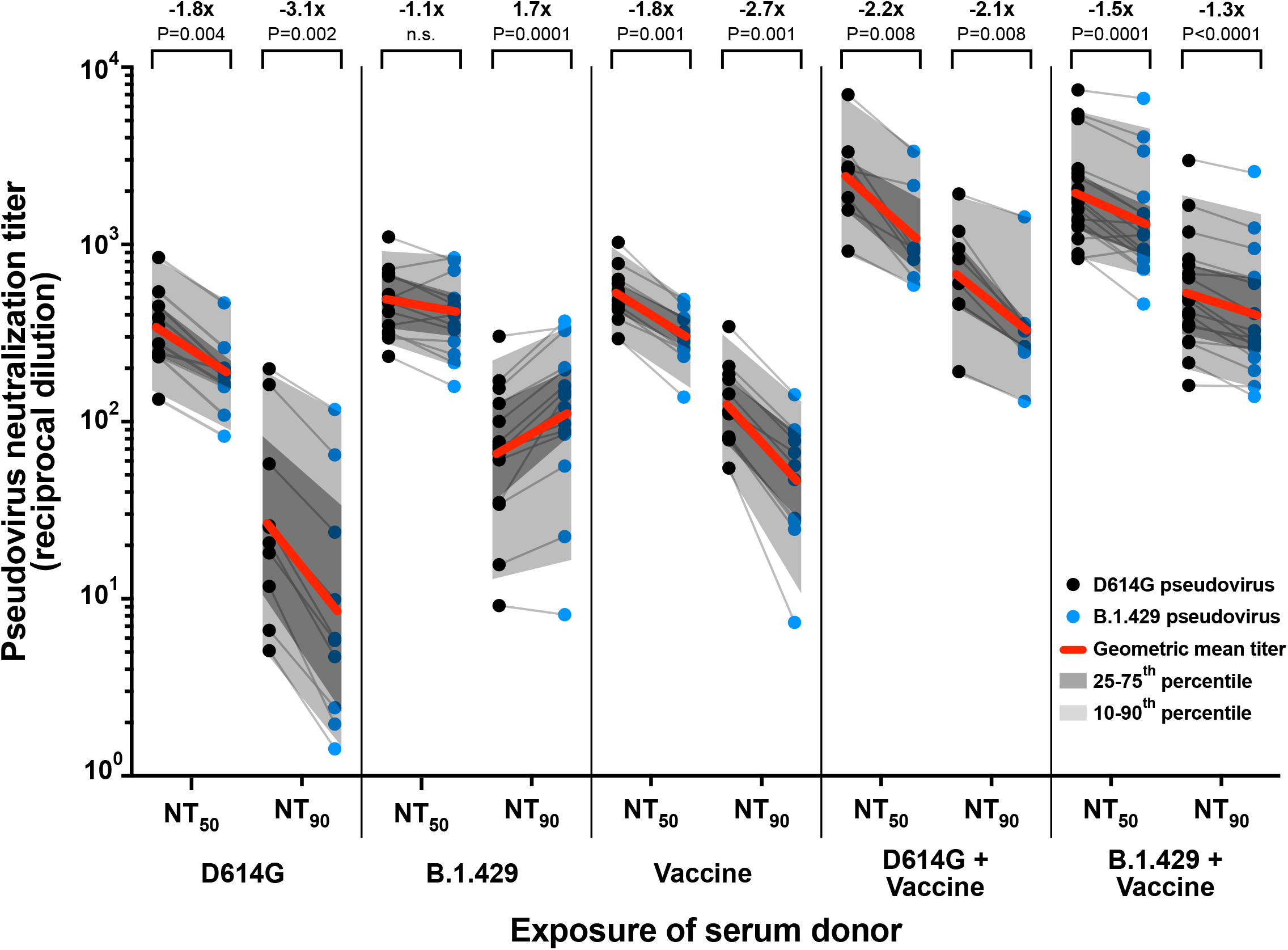
Neutralization of D614G and B.1.429 pseudoviruses by serum from individuals with different exposures. Plot of 50% and 90% pseudovirus neutralization titers (NT_50_ and NT_90_) of serum samples obtained from donors with the indicated infection and/or vaccination exposures. Grey lines connect neutralization titer values for D614G (black dots) and B.1.429 (blue dots) pseudoviruses within each individual serum sample. Geometric mean neutralization titers for each serum group are marked with red lines and fold-change in NT_50_ and NT_90_ between D614G and B.1.429 pseudoviruses is shown along with P-value. Dark grey shading marks the interquartile range of titer values in each group and light grey shading marks the 10^th^-90^th^ percentile of the range. P-values were calculated with a Wilcoxon matched-pairs signed-rank test.

We next investigated how exposure impacts neutralization specificity by crossing a panel of eight different spike variants against serum pools elicited by nine different prior exposures. (Figure 2; Table S1B). A range of reductions in neutralization titer relative to D614G pseudovirus were observed, with B.1.617.2 (Delta), B.1.351 (Beta), and B.1.1.529 (Omicron) exhibiting the greatest resistance to neutralization in serum from vaccinated or D614G-exposed individuals with up to 4-fold, 12-fold, and 65-fold reductions in NT_90_, respectively. However, for most variants, reductions in neutralization titer were considerably smaller or absent in serum from subjects previously exposed to a variant bearing some or all of the same spike mutations as the variant being tested. Specifically, prior exposure to the E484K mutation in the spike receptor binding domain (RBD) produced the greatest neutralization of four tested variants with mutations at the E484 position: B.1.617, P.1 (Gamma), P.2 (Zeta), and B.1.351 (Beta).

**Figure 2.**
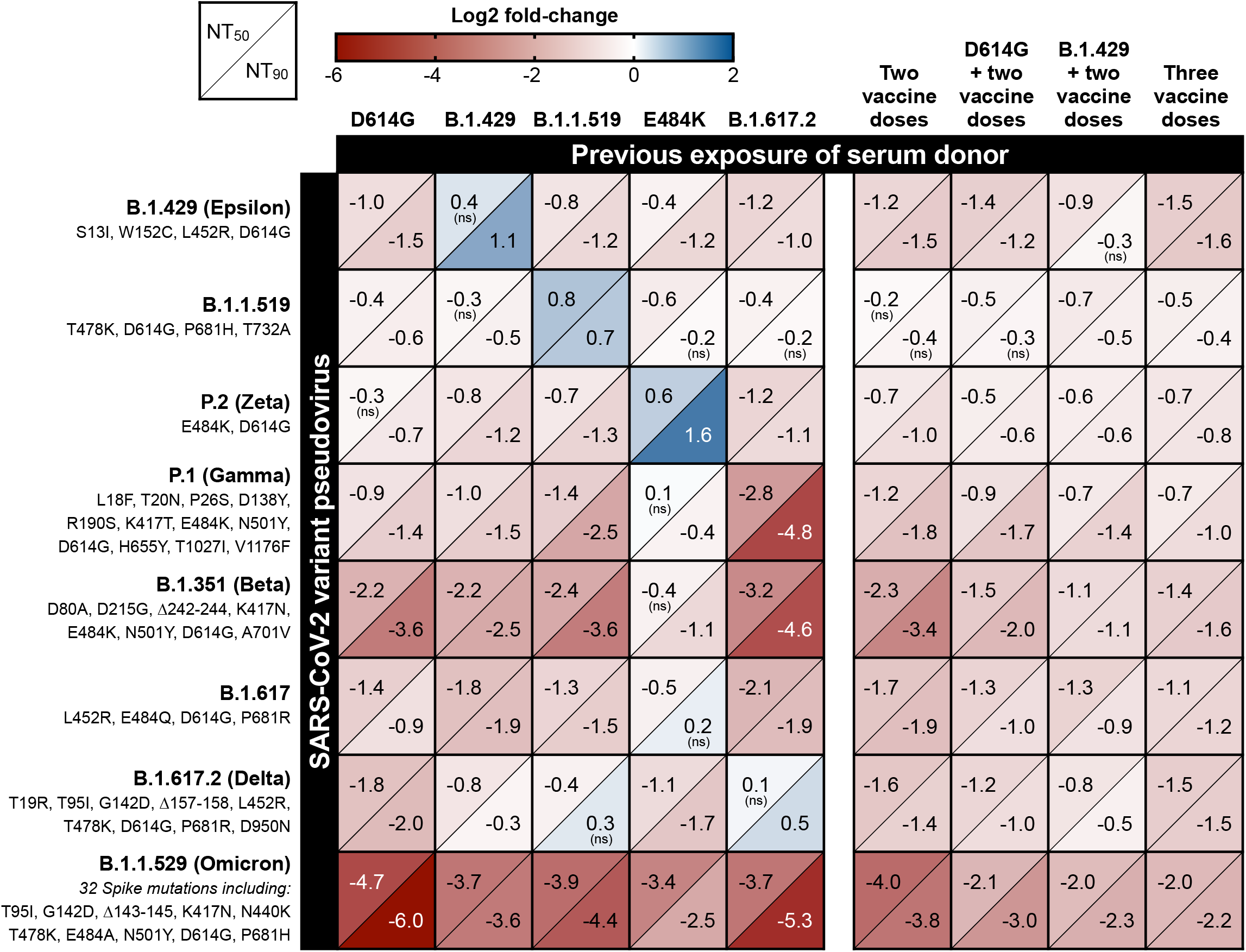
Change in variant pseudovirus neutralization titer relative to D614G. Matrix of normalized neutralization titers for seven different variant pseudoviruses (rows) neutralized by seven different pools of individual sera grouped by exposure (columns). Data is represented as a heat map of the Log2 fold-change in NT_50_ (top left of each box) and NT_90_ (bottom right of each box) of each variant relative to D614G pseudovirus. All serum samples were collected at least 14 days after the date of the subject’s positive COVID-19 test or date of most recent vaccine dose. All titer measurements are the mean of at least three independent experiments, each performed with two technical replicates. Positive Log2 fold-change (blue) indicates an increase in neutralization titer for that variant relative to D614G pseudovirus, while negative Log2 fold-change (red) indicates a decrease relative to D614G. Statistical significance was determined with unpaired t-tests. All values are statistically significant (P-value < 0.05) except where noted with “ns” to indicate the difference in variant neutralization titer is not significantly different from D614G pseudovirus neutralization titer in that serum pool.

Similarly, B.1.617.2 (Delta) was neutralized more effectively by serum elicited by partially homologous exposures B.1.1.519 and B.1.429 and was neutralized most effectively by serum elicited by fully homologous B.1.617.2 exposure. Conversely, in B.1.617.2-exposed serum we observed the least efficient neutralization of the highly divergent spike variants P.1 and B.1.351. Interestingly, although B.1.1.529 (Omicron) substantially escaped neutralization in all convalescent sera and serum from recipients of two vaccine doses, a much more modest 4 to 8-fold reduction in neutralization titer was observed in sera from individuals with previous infection plus vaccination or three vaccine doses.

## Discussion

In this study, we observe that vaccination and natural SARS-CoV-2 infection elicit neutralizing antibody responses that are most potent against variants that bear spike mutations present in the immunizing exposure. This trend is exemplified by variants with mutations at the spike E484 position, which were neutralized more effectively by E484K-exposed serum than other serum types. Importantly, we also show that B.1.617.2 (Delta) is neutralized more effectively by serum elicited by prior exposure to three different variants — B.1.429, B.1.1.519, and B.1.617.2 — which have separate sets of spike mutations partially or fully overlapping with mutations in B.1.617.2. These effects are presumably due to the shared L452R RBD mutation in B.1.429 and B.1.617.2, and the shared T478K RBD mutation and P681 furin cleavage site mutation found in both B.1.1.519 and B.1.617.2. The poor neutralization of P.1 and B.1.351 by Delta-exposed serum further reinforces the notion that cross-neutralization is heavily impacted by antigenic distance between variants [12]. Together, these results demonstrate that serum neutralization specificity is strongest against variants fully homologous to the exposure, but even single shared spike mutations, particularly those in highly antigenic regions such as the RBD, can enhance cross-neutralization as supported in other studies [3,6,13].

This study also demonstrates the effect of serial exposure to repeated or novel versions of spike on neutralizing antibody response. Infection with B.1.429 (Epsilon) followed by vaccination led to greater cross-neutralization of B.1.429 and B.1.617.2 (Delta) compared to vaccination alone or D614G infection plus vaccination, supporting the notion that exposure to multiple spike variants expands neutralization breadth. Repeated immunizing exposures from infection plus vaccination or booster vaccination led to both an overall increase in neutralization titers and generally broadened neutralization specificity, particularly towards B.1.1.529 (Omicron), which was neutralized most effectively by serum from recipients of three vaccine doses. A limitation of this study is the relatively small number of serum samples, however the shift in neutralization titer between D614G and variant pseudoviruses shows strong consistency between samples.

These serology data leverage human exposures to an array of naturally occurring spike mutations, including those relevant to the globally dominant B.1.617.2 and recently ascendant B.1.1.529 variants, providing a real-world complement to previous animal studies investigating heterologous boosting or multivalent vaccination strategies [14,15]. Our findings suggest that immunity acquired through natural infection will differ significantly between populations in different regions of the world due to highly variable prevalence of different SARS-CoV-2 variants throughout the course of the ongoing pandemic. These results also reinforce the urgent need for widespread booster vaccination and contribute additional evidence suggesting that heterologous or multivalent boosting strategies may be important and effective measures to address newly emergent variants such as the highly immune evasive B.1.1.529 (Omicron). Future studies investigating immune responses to additional emerging variants in vaccinated and unvaccinated individuals will contribute to identifying spike antigen versions that elicit broadly neutralizing antibody responses.

## Data Availability

Relevant viral genome sequences have been deposited in GISAID.
https://www.gisaid.org

## Acknowledgements

We would like to thank Dr. Chuka Didigu, Dorothy Park CRNA, Salu Ribeiro, and Bay Area Phlebotomy and Laboratory services for performing blood draws of study subjects. We thank Dr. Andreas Puschnik for providing the engineered cell line used in this study. We thank Susana Elledge and Dr. James Wells for providing reagents and advice on antibody assays. We thank Drs. Peter Kim, Don Ganem, Sandy Schmidt, and Cori Bargmann for technical assistance and discussion.

